# Impact of valganciclovir therapy on severe IRIS-Kaposi Sarcoma mortality: an open-label, parallel, randomized controlled-trial

**DOI:** 10.1101/2021.10.24.21265453

**Authors:** Patricia Volkow, Leslie Chávez Galán, Lucero Ramón-Luing, Judith Cruz-Velazquez, Patricia Cornejo-Juárez, Isabel Sada-Ovalle, Rogelio Pérez-Padilla, Islas-Muñoz Beda, on behalf of the Kaposi Sarcoma Study Group.

## Abstract

**Introduction:** High HHV-8 viral load (VL) in Kaposi Sarcoma (KS) has been associated with severe Immune reconstitution inflammatory syndrome (S-IRIS-KS), which can occur after initiating cART, and is linked with high mortality particularly in patients with pulmonary involvement. We investigate if valganciclovir initiated before cART decreases HHV-8 VL and assess if it reduces the incidence of S-IRIS-KS and its attributable mortality.

**Methods:** Open-label parallel-group randomized clinical trial in AIDS cART naïve patients with disseminated KS (DKS) as defined by at least two of the following: pulmonary, lymph-node or gastrointestinal involvement, lymphedema, or ≥30 skin lesions. In the experimental group (EG), patients were randomized to valganciclovir 900 mg BID four weeks before cART and continued until week-48; in the control group (CG), cART was initiated on week-0. Non-severe-IRIS-KS was defined as: increase in number of lesions plus ≥one log10 HIV-VL decrease or ≥50 cells/mm3 increase or ≥2-fold rise in baseline CD4+cells. S-IRIS-KS was defined as abrupt clinical worsening of KS lesions and/or fever after ruling out another infection following cART initiation, and at least three of the following: thrombocytopenia, anemia, hyponatremia, or hypoalbuminemia.

**Results:** 40 patients were randomized and 37 completed the study. In the ITT analysis, the overall mortality did not differ between groups. In the per-protocol analyses, the difference showed a trend for higher S-IRIS-KS mortality in the CG 3/19 (15.7%), compared to EG 0/18 (p=0.07). The incidence of S-IRIS KS was significantly lower in the EG; two patients, one each had S-IRIS-KS episode (0.038 per 100 patient-days) compared to CG group, four patients developed 12 S-IRIS-KS episodes (0.21 per 100 patient-days); incidence rate of 0.09 (95% CI 0.02-0.5 p=0.006). Mortality in patients with pulmonary KS was significantly lower in EG, 3/4 in CG vs 0/5 in EG. S-IRIS-KS was associated with higher HHV-8-VL; IL6 and CRP; valganciclovir was protective. Of survivors at week 48, 82% achieved >80% remission. No difference was found between groups in the number of non-S-IRIS-KS events.

**Conclusions:** Valganciclovir significantly reduced the episodes of S-IRIS-KS although attributable KS mortality was lower in the EG the difference was not significant (p=0.07). Mortality was significantly lower in EG patients with pulmonary KS.

NIH Clinical Trails ID NCT03296553.

## Introduction

Kaposi Sarcoma (KS) and *Pneumocystis jirovecci* pneumonia heralded the beginning of the HIV/AIDS pandemic [1,2]. Prior to the era of combined antiretroviral therapy (cART), over 30% of AIDS patients developed KS [3,4] which was considered a second epidemic among men that had sex with men (MSM) [5,6]. The incidence of KS dramatically decreased with the advent of cART, though KS attributable mortality is still high in the first months after cART initiation compared to AIDS patients without KS [7–10]. The high mortality observed shortly after cART initiation is mainly attributable to severe Immune Recovery Inflammatory Syndrome KS associated (S-IRIS-KS), which presents as an abrupt clinical worsening of KS alongside severe thrombocytopenia and other laboratory abnormalities [11–13] The clinical presentation of S-IRIS-KS shares similarities with Kaposi Sarcoma inflammatory Syndrome (KICS) but different from the later it has an abrupt onset, it can have recurring episodes and can responds to vincristine/bleomycin administration. Mortality from S-IRIS-KS may be as high as 25-40%, [10,11] which highlights the urgency to identify therapeutic interventions to prevent or reduce the severity of S-IRIS-KS.

High HHV-8 VL levels are associated with disease severity and mortality [20,21] and, are considered a risk factor for the development of IRIS-KS [11,15,16]. Thus targeting HHV-8 to diminish VL with drugs such as Ganciclovir and Foscarnet, which have shown *in vitro* and *in vivo* activity against HHV-8 [17–19] could be valuable for preventing IRIS-KS or for reducing its mortality. Ganciclovir, prior to the cART era, was used for secondary prophylaxis against Cytomegalovirus (CMV) end-organ disease and was associated with decreased KS incidence [20,21]. A clinical trial showed that valganciclovir reduced oropharyngeal HHV-8 replication [22].

Here we performed a randomized clinical trial to test the hypothesis that valganciclovir (ganciclovir prodrug) treatment prior to cART initiation, in patients with disseminated Kaposi sarcoma (DKS), would reduce HHV-8 VL and the occurrence of S-IRIS-KS and its associated mortality. We also evaluated whether the presence of coinfections, serum cytokines levels (IL6, IL10, TNF and IFN-⁤,) and C-reactive Protein (CRP) correlated with S-IRIS-KS occurrence.

### Patients and Methods

The protocol was approved by the Institutional Ethics Committee (015/031/INI) (CEI/950/15) in accordance with the Declaration of Helsinki and US Federal Policy for the Protection of Human Subjects and registered at NIH Clinical Trails ID NCT03296553. All participants signed written informed consent.

We conducted an open-label parallel-group randomized clinical trial of patients with DKS (all within the context of T_1_I_1_S_1_ Poor Prognosis according to the European Consensus-Based Interdisciplinary Guideline [23] The report of the trial follows the CONSORT guidelines [24].

Patients were recruited from October 21, 2015, to September 4, 2018. The last follow-up visit was on August 27, 2019.

Candidates had an initial thorough clinical evaluation including a work-up to diagnose coinfections and rule out other neoplasms that comprised: ophthalmologic evaluation, computed tomography (CT) scan (neck, thorax and abdomen), bone marrow culture with bone biopsy, Hepatitis B Virus (HBV) and Hepatitis C Virus (HCV) serology, venereal disease research laboratory (VDRL) test and if indicated lumbar puncture to rule-out neurosyphilis, Histoplasma urinary antigen and serology and GenXpert MTB/RIF test. Upper gastrointestinal tract (GIT) endoscopy with biopsy of lesions; colonoscopy was performed only on patients with diarrhea or lower-GIT bleeding. Biopsies of enlarged lymph-nodes were processed for culture and histopathological analysis. If indicated, we performed bronchoscopy with bronchoalveolar lavage (BAL), thoracic Gallium Scan or PET-FDG scan.

Inclusion criteria: Patients >18 years old, HIV+ naïve to cART with DKS, able and willing to provide written informed consent.

DKS was defined as the presence of: KS pulmonary disease and/or ≥30 KS skin lesions, with or without lymphedema, and/or lymph-node involvement, and/or GIT KS involvement (biopsy proven at least in one site).

Exclusion criteria: Another concomitant malignancy, Multicentric Castleman Disease (MCD), steroid treatment two months prior to screening, active HBV or HCV or CMV end organ disease, or severely ill patients with APACHE II score >15.

#### Randomization

Patients were randomized by blocks of ten; the assigned group written in closed envelopes: either to the Control Group (CG) to start cART immediately according to current Mexican Guidelines [25]; or to the Experimental Group (EG) to receive valganciclovir 900 mg twice daily for 48 weeks and initiate cART at week 4 after randomization.

#### Procedures

Patients who at the time of diagnoses presented with extensive KS involvement or bulky oropharyngeal lesions were treated with vincristine 2 mg (reduced to 1 mg if albumin was <3 g/dL) plus bleomicyn 15 UI prior to cART, or at follow-up in case of S-IRIS-KS or persistence of extensive KS lesions.

Visits were scheduled at baseline, week 1, 2, 4, 8, 12, 16, 24, and 48. At each visit an Infectious Diseases specialist performed a clinical evaluation and skin lesions were photographed. Blood samples were obtained for: WBC count, complete blood chemistry, urine analysis, CRP, D-Dimer, HIV VL with Abbott Real time, HHV-8, CMV and Epstein-Barr Virus (EBV) VL ELITe MGB KIT by ELITe InGenius Software, CD4+ and CD8+ cells count and percentage, (flow cytometry, Facs Canto II, Becton Dickinson) CD4/CD8 ratio and plasma levels of interleukin 6, 10 (IL-6, IL-10), tumor necrosis factor (TNF) and interferon gamma (IFN-γ) were measured using a sandwich-type immunoassay, ELISA (Biolegend). Serology for syphilis, HBV and HCV was repeated at week 24 and 48. If patients had an exacerbation of KS outside the scheduled visit they were re-evaluated with laboratory tests and image studies.

#### Severity Criteria

For the purposes of this trial, non-severe-IRIS-KS was considered in patients with increase in the number of KS lesions plus ≥one log10 of HIV-1 RNA VL decrease or ≥50 cells/mm^3^ or ≥2-fold from baseline CD4+ cells increase (26) S-IRIS-KS defined as an abrupt clinical deterioration after cART initiation alongside the presence of at least two clinical and at least three laboratory criteria.

Clinical criteria: 1) Fever (no identified concomitant infection), 2) increase in the size or number of KS lesions, 3) exacerbation of lymphedema, 4) appearance or increase of otherwise unexplained lung opacities on the chest images with a negative Gallium-Scan negative and 4) appearance or increase of pleural effusion.

Laboratory criteria: 1) Thrombocytopenia <100,000 platelets/ml; 2) Anemia (decrease of at least 1 g/dl from previous measure and no obvious bleeding), 3) Hyponatremia <135 mEq/L and 4) Hypoalbuminemia <3.5 g/dL. This is the clinical picture associated to death.

### Outcomes

The main outcome was overall mortality, and KS attributable mortality at 48 weeks. Secondary outcomes were the number of S-SIRI-KS events and the number of patients with a least one severe event as well as Pulmonary KS mortality.

Improvement of KS lesions, and KS remission assessed by comparing the size and number of KS lesions from baseline to 48-week visit and lung CT-scan for cases with pulmonary involvement.

#### Statistical analysis

Sample size was calculated for a study power of 80% and an alpha of 0.05, with an estimated event rate (S-IRIS-KS) in the control group of 40%, and in the treated group of 5%. The number estimated in each group was 19, for a total sample of 38 patients.

Descriptive statistics were performed, including the number of deaths, and episodes of S-IRIS-KS, in the CG and EG groups.

Comparisons between EG and CG were performed, as intention-to-treat (ITT), and as per-protocol analysis, for the primary outcome of mortality attributed to S-IRIS-KS using Proportional-Hazard Cox models.

, We also compared the impact of treatment on repeated episodes of S-IRIS-KS by multivariate Poisson model and by the number of participants with at least one S-IRIS-KS event, as repeated events can occur in the same patient. In addition we compared mortality in EG and CG in patients with pulmonary KS involvement utilizing the Fisher ’s exact test.

Statistical models were adjusted by baseline CD4+, CD4+/CD8+ cell ratio, HHV-8 and HIV VL, plasmatic IL-6, CRP and by the presence of concomitant coinfections. Analyses were performed using Stata Statistical program.

## Results

A total of 91 male patients were screened, 40 of them were randomized Figure 1; between 10/2015 to 09/2018, all MSM, with a median age of 31 years (IQR 26-36), all Hispanic.

**Figure 1.**
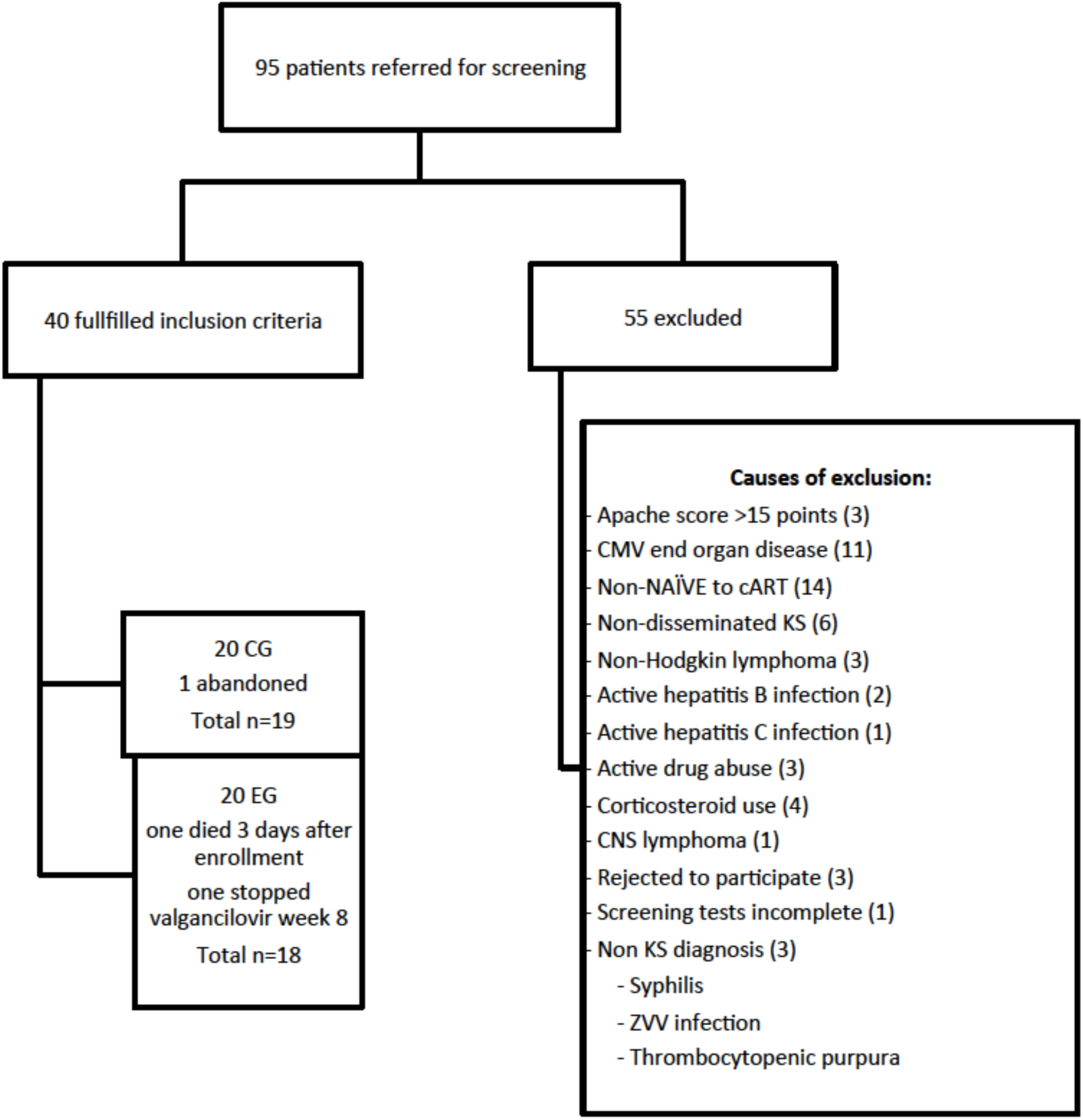
Screening diagram.

### ITT analysis

In the ITT analysis comparing all forty randomized patients the overall mortality did not differ between groups, three died in each group. The prevalence of syphilis, mycosis, mycobacterial and parasitic infections as well as laboratory parameters and cytokines levels did not differ between groups, except for IL-6, which was significantly higher in EG (Table 1). Thirty-five (87%) patients had HHV-8 viremia at baseline alone or in combination with EBV or CMV (19 EG and 16 CG). Nine (22.5%) patients had pulmonary KS involvement CG= four (20%); EG=five (25%). Eleven patients (27.5%) were diagnosed with HIV-polymorphic lymphoproliferative disorder [27].

**Table 1.**
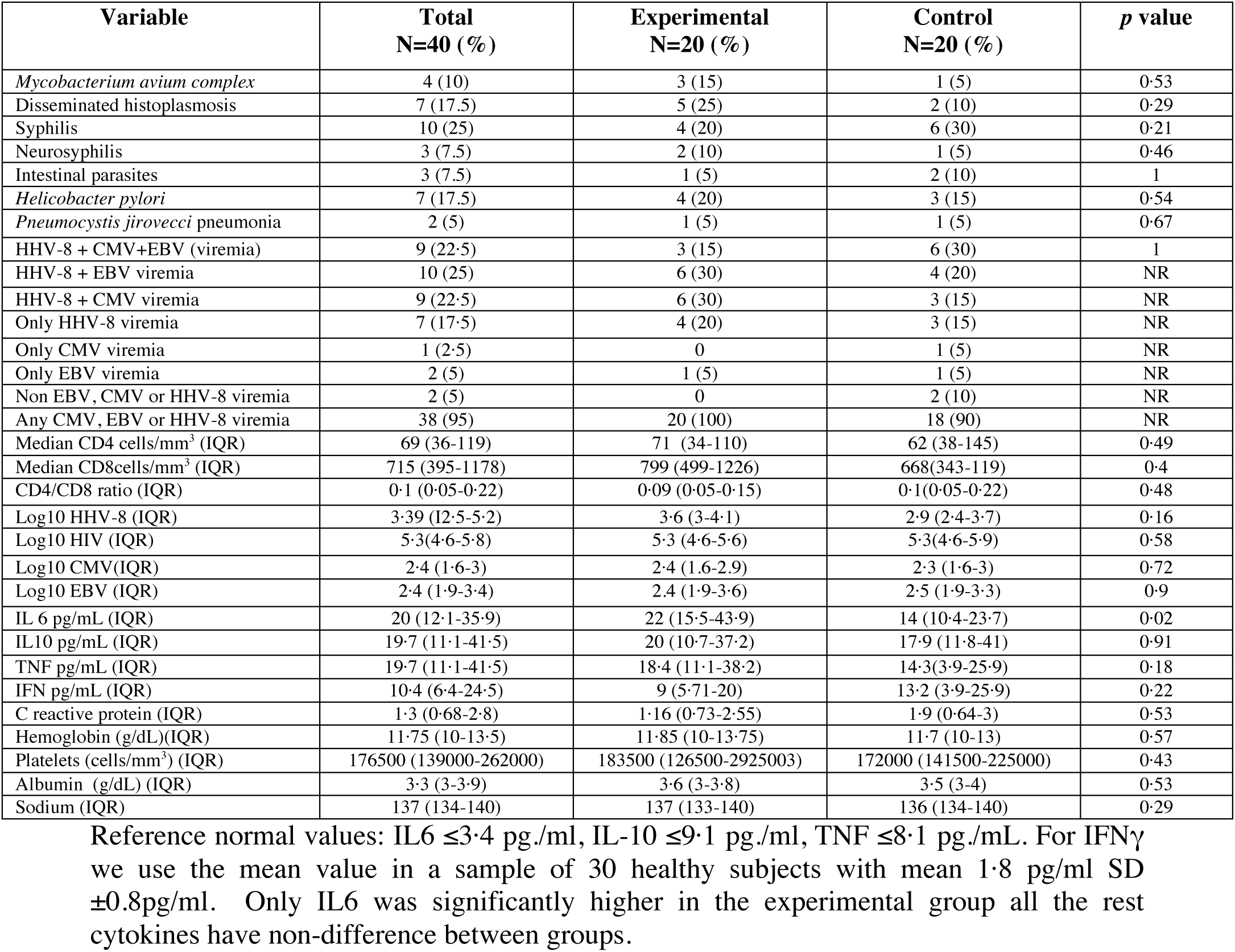
Prevalence of coinfections and Human Herpes virus 8 (HHV-8), Cytomegalovirus (CMV) and Epstein Barr virus (EBV) viremia, laboratory parameters and cytokines in the cohort of patients comparing experimental and control groups.

In the CG group all deaths were attributable to S-IRIS-KS, occurring 70, 88 and 99 days after randomization. In the EG group, one death occurred three days after enrollment with septic shock, one 89 days post randomization from an opioid overdose, and one 325 days post randomization due to H1NI influenza pneumonia (Table 2).

**Table 2:** Outcomes of the Trial: Mortality and Severe-IRIS-KS Events.

| Outcomes | Treatment Groups |  |  | Crude | Adjusted |
| --- | --- | --- | --- | --- | --- |
|  | Experimental<br>n=18 | Control<br>n=19 | *p* value | HR or IRR (95% CI) | HR or IRR (95% CI) |
| Primary outcome |  |  |  |  |  |
| Deaths at 48 weeks | 3/20 | 3/20 | 1 | 1.3 (0.25-6.8) *p*=0.8* | 0.45 (0.07-3.0) *p*=0.4* |
| KS-attributable deaths at 48 weeks (per protocol analysis) | 0/18 | 3/19 | 0.08 | *p*=0.09 log rank test | - |
| KS-attributable deaths at 48 weeks (ITT analysis) | 0/20 | 3/20 | 0.07 | *p*=0.09 Log rank test | - |
| Secondary outcome |  |  |  |  |  |
| Total number of S-IRIS-KS events in the group (incidence rate per 100 patient-days) | 2 (0.038) | 12 (0.21) | 0.024 | IRR 0.18 (0.04-0.8, P=0.024)** | IRR 0.09 (0.02-0.5, P=0.006)** |
| Participants with at least one S-IRIS-KS event | 2/18 | 4/19 | 0.71 | 0.53 (0.1-2.9 P=0.47)** | 0.28(0.04-2.1, P=0.21)** |

*p* with Chi-square test. \*HR= hazard ratio by Cox proportional hazard model. \*\*IRR=incidence rate ratio by a Poisson regression model. Models were adjusted by baseline CD4+ T cell count, IL-6 levels, log-viral load of HHV-8, presence of coinfections (yes or no) and presence of coinfections not curable in a short time. NR: not realized.

In the per-protocol analysis we included 37 patients, 19 in the CG (one patient abandoned the protocol on week 12) and 18 in the EG (one patient died three days after randomization and another stopped valganciclovir on week-8. Four patients developed 12 episodes of S-IRIS-KS in the CG Incidence Rate (IR) (0.21 per 100 patient-days) and two patients developed one episode each in the EG (IR 0.038 per 100 patient-days, *p* = 0.024). The multivariate Poisson model showed that higher HHV8-VL, IL6 and CRP increased the risk to develop S-IRIS-KS and valganciclovir use significantly decreased the risk of S-IRIS-KS occurrence (Table 3).

**Table 3.** Multivariate analyses for Severe-Immune Reconstitution Inflammatory Syndrome associated to Kaposi Sarcoma (IRIS-KS).

| S-IRIS-KS events | IRR | P>z | [95% Conf. Interval] |  |
| --- | --- | --- | --- | --- |
| Baseline CD4+ cells count | 1.36 | 0.17 | -0.002 | 0.01 |
| Valganciclovir therapy | -3.39 | 0.001 | -5.76 | -1.54 |
| Baseline HHV-8 viral load | 2.65 | 0.008 | 7.14 | 4.77 |
| C reactive protein | 2.51 | 0.01 | 0.47 | 0.38 |
| Baseline IL-6 | 3.27 | 0.001 | 0.44 | 0.17 |

Valganciclovir did not significantly reduce the number of patients with at least one S-IRIS-KS episode (adjusted IRR 0.28, 95%CI 0.04, 2.1, p= 0.21) but it significantly reduced the total number of S-IRIS-KS events (adjusted IRR= 0.09, 95%CI 0.02-0.5 p=0.006). The risk of death in the Cox regression analysis was increased by 12 (95%CI 1-145) when HHV-8 VL at baseline was >5000 copies/ml (Table 3).

Of the nine patients with pulmonary KS, three of the four in the CG group (75%) died of S-IRIS-KS, whereas none of the five patients in the EG died of S-IRIS-KS (Fisher ’s exact test p=0.048). No difference was found in the occurrence of non S-IRIS-KS events; eight patients in each group did not develop any IRIS event, EG=7 patients developed 15 and CG=8 patients developed 17 non-S-IRIS-KS events.

There were no differences in the number of patients receiving or the number of cycles of vincristine/bleomycin administered. Fourteen (37.8%) patients did not receive chemotherapy (CG=7; EG=7), eight received one cycle (CG=four; EG=4), eight two cycles (CG=3; EG=5), three patients three cycles (CG=2; EG=1), one patient four cycles CG and three patients received five cycles (CG=2; EG=1).

At week-48 follow-up 33 patients were alive and one CG was lost to follow-up. There were no group differences in remission rates; 12 (36.3%) achieved complete remission (six in each group); 15 (45.4.7%) achieved 80-95% remission (CG=seven, EG=eight); and six (18%) 50-70% remission (three in each group). We did not document any relapses or non-responders.

### Comparison baseline *vs*. week-48 measurements

In survivors most measurements improved from baseline to follow-up at week-48; the rate of improvement did not differ between groups.

The median CD4+ cells count was 69 cells/mL (IQR 36-119) at baseline and 179 cells/mL (IQR 132-516) at week-48. The median CD8+ at baseline was 715 cells/mL (IQR 395-1178) and 639 cells/mL (IQR 639-673) at week-48. The median CD4+/CD8+ ratio was 0.1 (IQR 0.05-0.22) at baseline and 0.34 (0.16-0.48) at week-48.

At baseline, the median of IL-6 was 20 pg/ml (IQR 12.1-35.9) (reference value (RV) of <3.4 pg/ml); higher for EG 22 pg/ml; (IQR 15.5-43.9) than for CG 14 pg/ml, (IQR10.4-23.7); p=0.02) Figure 2. Baseline levels for IL-10 19.7 pg/ml, (IQR 11.1-41.5; RV <9.1 pg/ml); IFNγ 10.4 pg./ml, (IQR 6.4-24.5, RV=1.8pg/ml SD± 0.8pg, RV 4 pg/ml [28] and TNF 19.7 pg/ml, (IQR 11.1-41.5, RV = 8.1 pg/ml) did not differ between groups.

**Figure 2.**
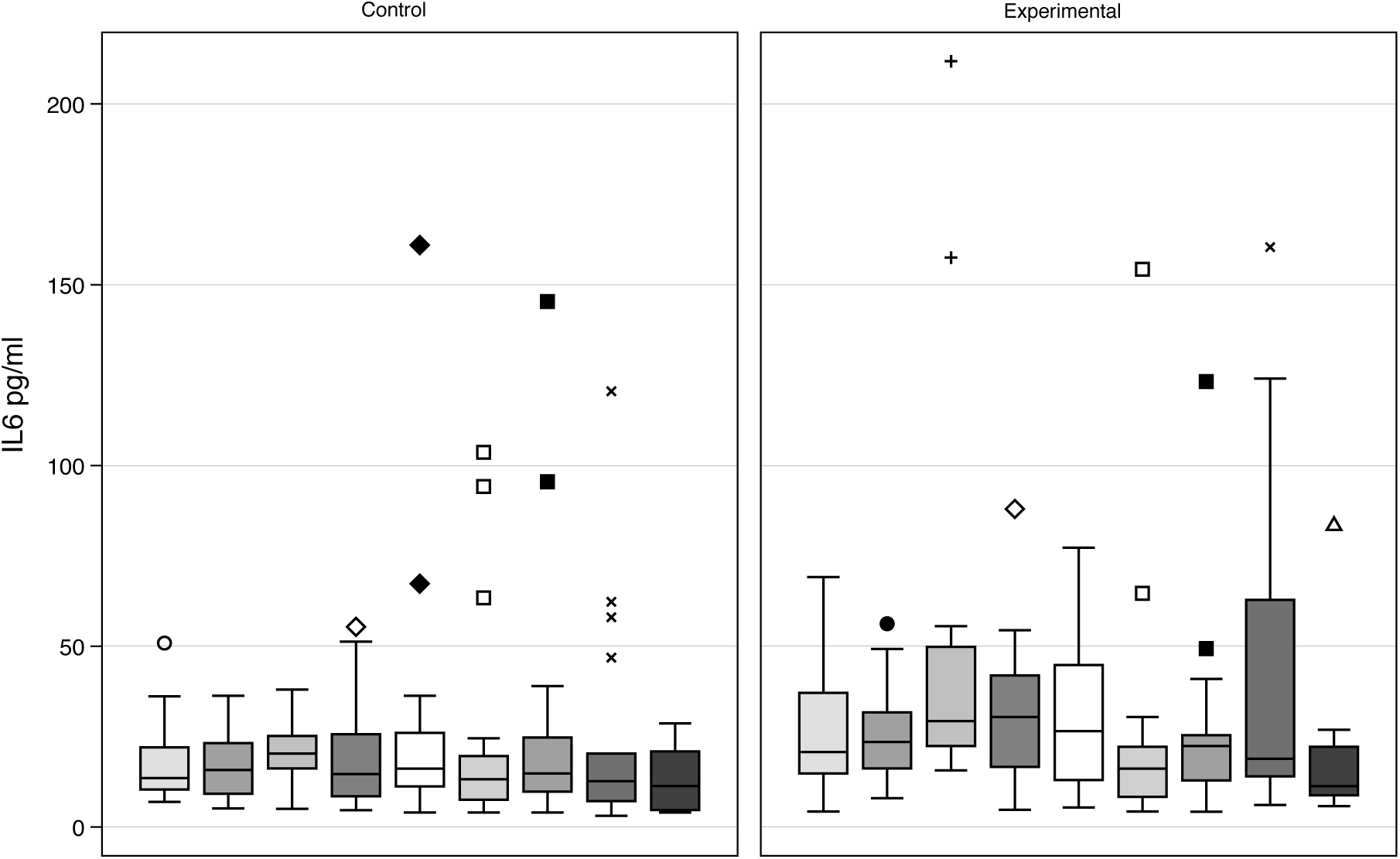
IL6 levels by group by protocol scheduled visit.

At week-48, IL6 levels for those who survived and were not lost to follow-up (n=33) were 11.3 pg/ml (IQR 7.4-21), IL10 13.8 (IQR 7.3-36), IFNγ 10.7 pg/ml (IQR 5.6-21.2), and TNF 13.05 pg/ml (IQR 7.7-21.2) there were no differences between groups.

The median CRP at baseline was 1.3 (IQR 0.68-2.28) and did not differ between groups (Table 1).

At week-48, 23 patients (65%) had undetectable HHV-8 VL, 13 in EG and ten CG. Three had detectable HHV-8 VL in EG (highest measurement was 929 copies/ml) and six in CG (three of them with HHV-8 VL >10000 copies/ml) Figure 3. Thirty-one patients (93.9%) had undetectable HIV VL, (<45 copies/ml) and two had HIV viremia (6.1%), both with adherence problems.

**Figure 3.**
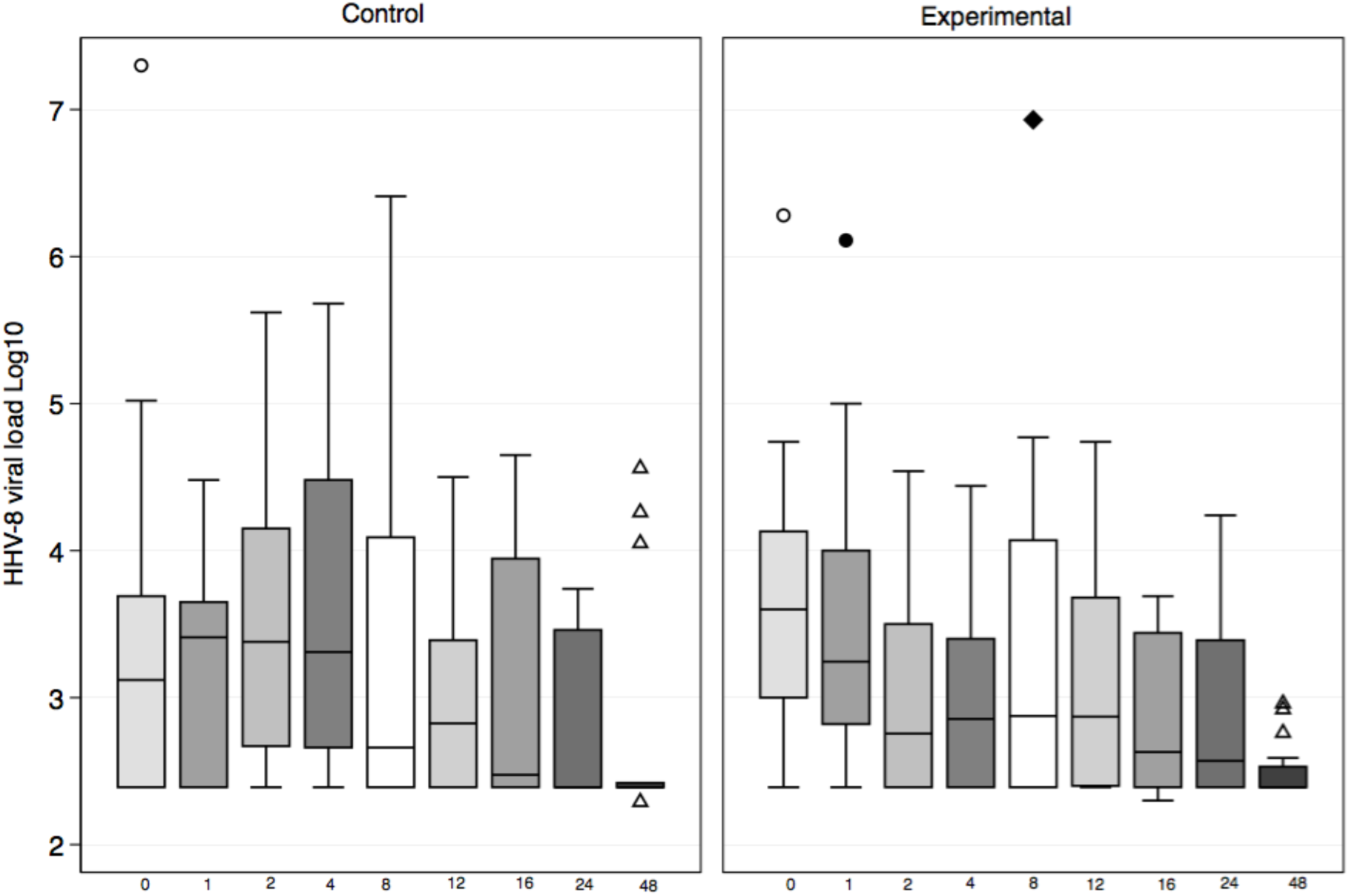
Log10 HHV-8 VL by group by protocol scheduled visit.

### Cytokines according to S-IRIS-KS

This analysis was done with the patients included in the ITT. The median baseline IL-6 levels in the patients who developed S-IRIS-KS was 36 pg/ml (IQR 16.1-50) and significantly higher than in patients without S-IRIS-KS events 15.5 pg./ml (IQR 11.7-22) (*p*=0.018).

Median baseline levels were also higher for IL-10 76.8 pg/ml, (IQR 21-1075) in patients who developed S-IRIS-KS versus those who did not 15.1 pg/ml (IQR 9.9-32) (p=0.01). There were no differences in baseline TNF levels among patients who developed S-IRIS-KS (16.7. pg/ml IQR 3.4-16.7) compared to those who did not (18.07 pg/ml IQR 6.3-31.1) (p=0.85) nor were there differences for IFNγ; the median for those who developed S-IRIS-KS was 15.9 pg./ml (IQR 8-82.5) and 9.3 pg/ml for those who did not (IQR 5.8-18.7) (p=0.1).

CRP among patients that developed S-IRIS-KS was 3.4 (IQR 3.3-10.1) significantly higher than among those who did not 1.1 (IQR 0.49-1.9) (p=0.001).

Figure 4 depicts the CD4+ cell levels, HHV-8 and HIV viral load for each visit for one patient from the CG and one from the EG, both with pulmonary involvement. The S-IRIS-K events are shown along with the outcomes.

**Figure 4.**
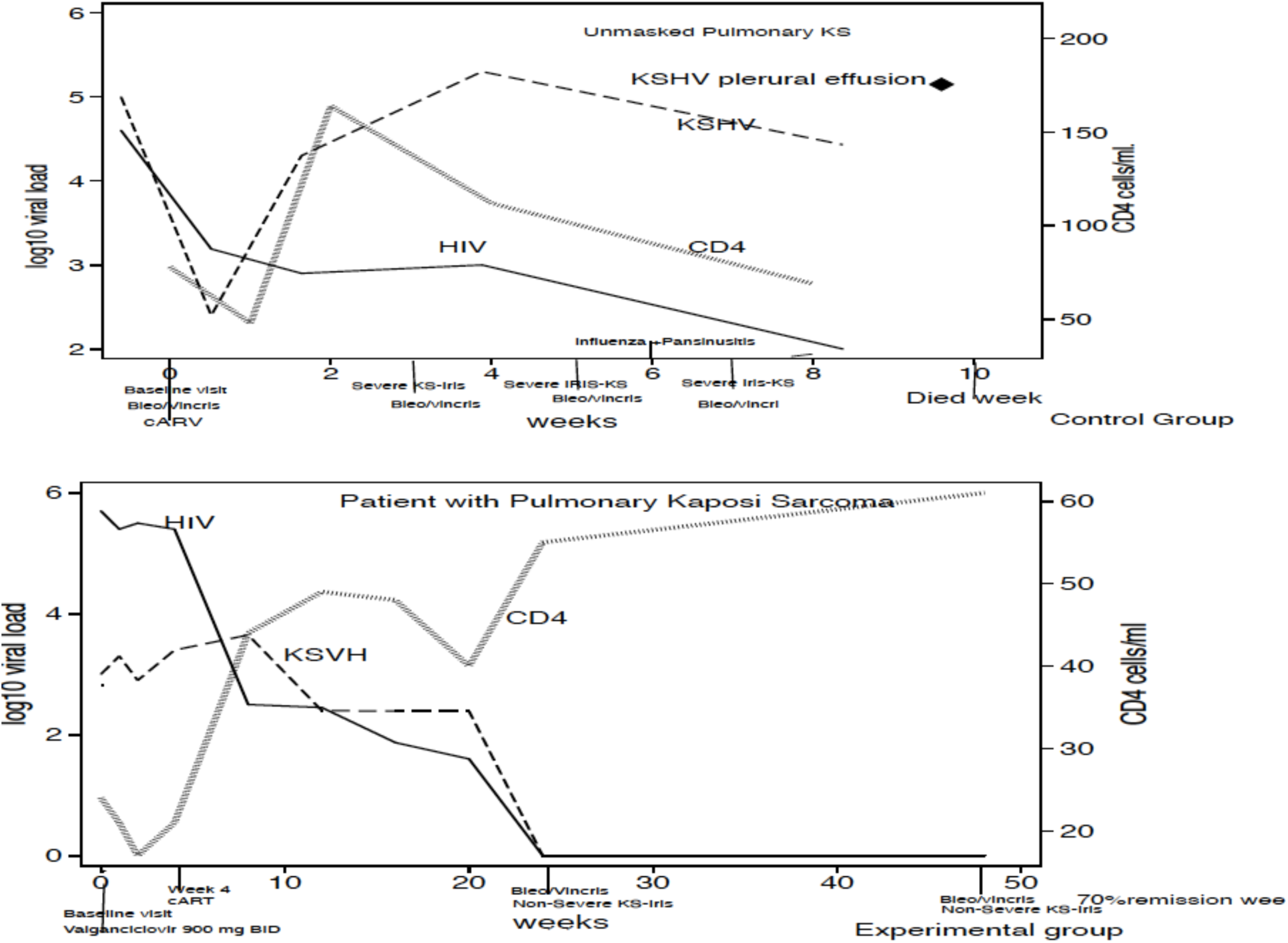
Clinical evolution, CD4+ cell count, HIV and HHV-8 viral load for each visit in a patient from the control group (upper panel) and in a patient from the experimental group (lower panel). The patients in the control group developed S-IRIS-KS and died and the patient in the experimental group had pulmonary KS and had an uneventful evolution.

Figure 5 depicts these measures in the patient who stopped valganciclovir on week-8 and resumed it on week-24. This illustrative case shows that the patient developed S-IRIS-KS after valganciclovir discontinuation, when HHV-8 VL rebounded and CD4+ cells increased, and when he resumed treatment, HHV-8 VL decreased and the S-IRIS-KS episode ceased, achieving 90% remission at week-48.

**Figure 5.**
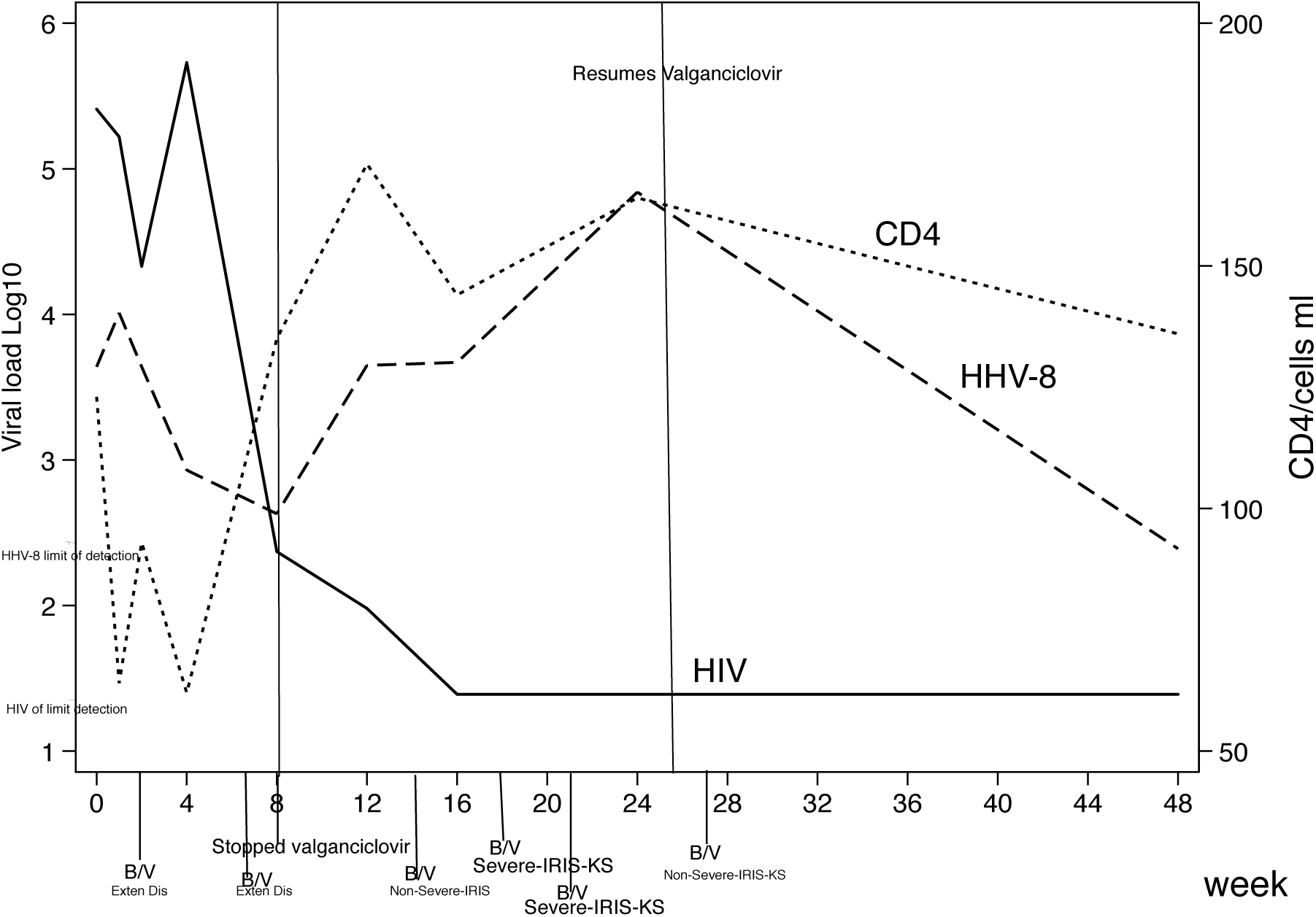
Patient in EG that interrupted valganciclovir treatment on week 8, his HHV-8 VL rebound and on week-21 developed S-IRIS-KS. Valganciclovir was re-started on week24; HHV-8 VL replication was finally suppressed. This subject was excluded in the per-protocol analysis.

### Adverse events

Complete adverse events are displayed in Table 4.

**Table 4.** Adverse events.

| Adverse Event | Total<br>N=37 | Experimental<br>N=18 | Control<br>N=19 |
| --- | --- | --- | --- |
| Severe neutropenia <500-100 cells/mm<sup>3</sup> | 5 | 3* | 2 |
| Mild neutropenia ≥500 cells/mm<sup>3</sup> | 3 | 2<sup>+</sup> | 1<sup>‡</sup> |
| Febrile Neutropenia | 1 | 1 | 0 |
| Cellulitis | 6 | 4 | 2 |
| Allergic Rash<sup>†</sup> | 8 | 4 | 4 |
| *C. difficile* infection & | 1 | 1 | 0 |
| Pneumonia | 2 | 2 | 0 |
| Multifocal leukoencephalopathy | 1 | 1 | 0 |
| UTI *E.coli* ESBL | 2 | 0 | 2 |
| Opioid Overdose | 1 | 1 | 0 |
| A H1N1 | 1 | 1 | 0 |
| Abdominal sub occlusion | 1 | 1** | 0 |
| Oral Mucosa dysplasia | 1 | 0 | 1 |
| Myositis | 1 | 0 | 1 § |
| Acute necrotizing ulcerative gingivitis | 1 | 1 | 0 |
| Lower limb thrombosis | 1 | 1 | 0 |
| Heaviness and mild pain in legs | 1 | 0 | 1 |
| Suicidal ideation | 1 | 1 | 0 |
| Neurologic disturbance | 2 | 1 | 1 |

\* All three patients had disseminated histoplasmosis.
‡ One patient in each group had *Mycobacterium Avium Complex* (MAC) infection.
† Four patients had rash attributed to efavirenz and four due to TMP/SMX.
& One patient in the experimental group presented *C. difficile* infection with one relapse.
\*\*Attributable to MAC immune reconstitution inflammatory syndrome with abdominal lymph node enlargement.
§ Attributed to Tenofovir.

Ten patients acquired a sexually transmitted disease (STD) during the time period of the study: eight syphilis, three at week 24 and five at week 48, one gonococcal pharyngitis and one pediculosis pubis.

The use of Filgastrim was significantly greater in EG OR 3.13 *p*=<0.005

## Discussion

In this randomized clinical trial we evaluated the effect of valganciclovir as antiviral for HHV8 on DKS patients who were cART naïve, with severe immunosuppression, comorbid coinfections and variable levels of HHV-8 viremia. Participants were randomized to receive valganciclovir four weeks prior to cART initiation and continued for 48 weeks (EG) or to control group (CG) who received cART upon randomization. Although we could not observe differences in mortality in the ITT analysis, we found a significant decrease in the occurrence of S-IRIS-KS (*p*=0.006) and a trend for lower KS attributable mortality (*p* =0.07) in the EG compared to the CG groups.

KS is an angioproliferative disease caused by HHV-8, mediated by cytokines in severely immunosuppressed individuals [29]. As all herpes viruses, HHV-8 has a latent and a lytic replication phase [30] and a genetic machinery that mimics human oncogenes [29]. In the latent phase only six genes are expressed, including ORF50, which codifies for P53 decreasing apoptosis and increasing the survival of infected cells and evades the host’s immune system allowing the virus to persist [31,32]. In the lytic phase all genes that promote replication are expressed [33] resulting in the release of progeny virions and death of the infected cells. They also promote cell division of infected cells, inhibit apoptosis, modulate inflammation, induce angiogenesis [34] though it does not produce cell immortalization [35]. KS is considered a multicentric, policlonal disease [4,36,37].

There are multiple stimuli that promote the lytic phase of the virus, including several inflammatory cytokines [38] like IL-6, oncostatin, alfa-TNF, platelet-derived growth factor (PDGF), vascular endothelial growth factor (VEGF) that can promote the Replication Transcription Activator (RTA) pathway, which is a potent lytic activator of HHV-8 [33]. The HIV-Tat protein induces expression of cytokines like N-kappa B, IFNγ and VEGF that also activate the lytic phase [39] and HIV itself results in immunodeficiency (a *syne qua none* factor) facilitating KS development [35,40,41]. There is evidence that HHV-8 replication correlates with extensive KS and disease progression [15,43,44].

Immunosuppression favors coinfections with opportunistic pathogens and the infectious processes promote production of inflammatory cytokines and stimulation of RTA and thus lytic HHV-8 replication. We found high levels at baseline of the four cytokines studied (IL-6, Il-10, IFNγ and TNF) (Table1). Both Il-6 and IL-10 were significantly higher among patients that developed S-IRIS-KS; whereas IFN-ϒ and TNF showed no differences. High IL-10 levels have been demonstrated in patients with DKS [45]. In this cohort we found a high prevalence of multiple infections including syphilis 25%, Histoplasmosis and *Helicobacter pylori infection* (17.5% each), MAC in 10%, parasites and a diverse combination of HHV-8, CMV and EBV viremia. In the early years of the AIDS epidemic, similar data were published in DKS case reports; and it was postulated that these coinfections particularly CMV and other STDs could participate in the pathogenesis of DKS [2,46]. Despite evidence of the high prevalence of coinfections in HIV patients [47] only opportunistic infections are addressed in KS guidelines, none of the other type of infections are mentioned [23,48].

Coinfections in DKS patients should be diagnosed and treated to avoid the vicious cycle of inflammatory cytokines production that promote lytic phase activation and thus HHV-8 replication and in consequence increase in HHV-8 VL [38].

We postulate that valganciclovir diminishes HHV-8 VL modulating inflammatory cytokine release when HIV VL diminishes and CD4+ cells begin to increase. Indeed during a S-IRIS-KS event, HHV-8 VL increases in parallel to CD4+ cells rise (see Figure 5) this is apparent in the patient who stopped valganciclovir while continuing to receive cART, at which point he developed an episode of S-IRIS-KS. Our study demonstrates that valganciclovir decreased S-IRIS–KS episodes and although we did not observe a significant decrease in mortality this probably reflects the small sample size of our study.

The use of low doses antimicrotubule agents like vincristine, that halt the cellular cycle, can impact the production and release of virions from HHV-8 infected cells and be clinically beneficial. Patients with extended disease or S-IRIS-KS were treated with 2 mg vincristine and 15UI bleomycin. This regime was well tolerated; it is not myelosuppressive and does not require concomitant administration of steroids, a recognized amplifier of KS proliferating signaling [45].

The delay in the initiation of cART while giving an anti-HHV8 agent has the same basis as when cART initiation is delayed in CMV chorioretinitis or cryptococcocal meningitis, [49– 52] and is to diminish the risk of IRIS development and its deleterious consequences to the patient’s health.

HHV-8 also denominated Kaposi sarcoma Herpes virus (KSHV) is recognized as the causing agent of KS, MCD, Pulmonary Effusion Lymphoma (PEL) and KICS [36,53]. Also recognized is the role of cytokine dysregulation particularly IL6 and IL10, on the pathogenesis of these diseases, which share similar clinical and laboratory abnormalities including thrombocytopenia, anemia, hipoalbuminemia, hiponatremia, edema and effusion. The criteria used to diagnosed S-IRIS-KS share similarities to KICS criteria [54,55], the main difference is the temporal relation to cART initiation, the abrupt initiation of symptoms (fever, increase in SK lesions, lymphedema or pleural effusion, rapid decrease in platelets, Hb, sodium and albumin) and the response to vincristine/bleomycin [56]. DKS typically has a high mortality particularly in patients with pulmonary involvement [57,58], we found a significant difference in mortality, in the CG 75% of patients with pulmonary KS died of S-IRIS-KS *vs*. none in the EG group.

The measurement of HHV-8 VL before initiating cART proved clinically useful in DKS patients. From our perspective, valganciclovir should be initiated in patients with pulmonary involvement and in patients with a VL of ≥ 5000 copies/ml as the latter had 12 times more risk of developing S-IRIS-KS. In patients with lower HHV-8 VL it should be tailored. We recommend repeating the HHV-8 VL four weeks after cART initiation and if there is an increase of one log on HHV-8 consider the use of valganciclovir.

The pathogenesis of DKS requires that cytokines activate RTA, which promotes the lytic phase of HHV-8 with the subsequent expression of oncogenes that then promote inflammation, proliferation and differentiation of spindle cells [59] and HHV-8 virions production, this in an environment of immunodeficiency. Thus it is important to tailor therapy to address the four elements of KS pathogenesis: (1) control of HHV-8 viremia with valganciclovir (or other anti-HHV8 active agent) [17,19], (2) diagnosis and treatment of co-infections, (3) suppression of HIV viremia to allow immune restoration [60] and (4) suppression of spindle-cell proliferation with antimicrotubule agents that do not exacerbate immunosuppression.

We acknowledge some limitations in our study. First, the sample size was small, and this may explain an absence of a statistically significant impact in KS attributable mortality. In estimating the sample size for the trial we overestimated the expected impact of valganciclovir treatment on mortality.

## Conclusions

Valganciclovir significantly reduced the episodes of S-IRIS-KS although attributable KS mortality tended to be lower in the EG the difference was not significant. In EG patients with pulmonary KS mortality was significantly lower.

## Data Availability

All data produced in the present study are available upon reasonable request to the authors

## Author’s contribution

All the authors contributed significantly to this work and participated in the drafting of the manuscript and have seen and approved the version submitted.

PVF designed the study, PVF and BIM participated in the care of patients and database, LChG and LRL processed and analyzed cytokines, JCV performed all herpes virus studies, and RPP, PVF, BIM and PCJ performed statistical analysis.

The Kaposi Sarcoma study group is formed by persons whose work was essential for the development of the study beside the main authors.

Carmen Lome-Maldonado performed the histopatologic evaluation of biopsies. Juan W Zinser help in the treatment with vincristine and bleomicyn.

Diana Vilar-Compte performed the sample size estimation and the randomization procedure.

Lucero González clinical nurse, obtained blood samples facilitated diagnostic and clinical studies, assisted during invasive procedures, obtained pictures and uploaded the data.

Ayumi Kawakami and Matilde Ruiz performed ophthalmologic evaluations.

## Acknowledgements

To patients. To their mothers and partners for their unconditional support.

And for all colleagues that help in any time the care of patients: Roberto Rodríguez.

Barbara Chavez Mazarri, Dra Brenda Gomez, Dra Isabel Sada, Dr. Daniel de la Rosa, Dr. Gustavo Rosales, Dr Erick Osorio, Dra Alexandra Martin-Onraet, Dra Carolina Pérez, QFB Consuelo Velazquez, SW Claudia Campos, Dr. Grimaldo Flavio, Dr. Arturo Martínez-Orozco.

